# RNA Polymerase II pausing temporally coordinates cell cycle progression and erythroid differentiation

**DOI:** 10.1101/2023.03.03.23286760

**Authors:** Danya J. Martell, Hope E. Merens, Claudia Fiorini, Alexis Caulier, Jacob C. Ulirsch, Robert Ietswaart, Karine Choquet, Giovanna Graziadei, Valentina Brancaleoni, Maria Domenica Cappellini, Caroline Scott, Nigel Roberts, Melanie Proven, Noémi BA Roy, Christian Babbs, Douglas R. Higgs, Vijay G. Sankaran, L. Stirling Churchman

## Abstract

The controlled release of promoter-proximal paused RNA polymerase II (Pol II) into productive elongation is a major step in gene regulation. However, functional analysis of Pol II pausing is difficult because factors that regulate pause release are almost all essential. In this study, we identified heterozygous loss-of-function mutations in *SUPT5H*, which encodes SPT5, in individuals with β-thalassemia unlinked to *HBB* mutations. During erythropoiesis in healthy human cells, cell cycle genes were highly paused at the transition from progenitors to precursors. When the pathogenic mutations were recapitulated by *SUPT5H* editing, Pol II pause release was globally disrupted, and the transition from progenitors to precursors was delayed, marked by a transient lag in erythroid-specific gene expression and cell cycle kinetics. Despite this delay, cells terminally differentiate, and cell cycle phase distributions normalize. Therefore, hindering pause release perturbs proliferation and differentiation dynamics at a key transition during erythropoiesis, revealing a role for Pol II pausing in the temporal coordination between the cell cycle and differentiation.

## Introduction

Cellular differentiation is a tightly regulated, multi-step process that requires several transitions in cell type–specific gene expression programs. Consequently, the precise regulation of transcription, a key step in controlling gene expression, is fundamental to cellular differentiation (Young 2011; Lee and Young 2013; Jonkers and Lis 2015). RNA polymerase II (Pol II) pauses 30–50 bp downstream of promoters, and its release into productive elongation is a highly regulated process (Chen, Smith, and Shilatifard 2018; Mayer, Landry, and Churchman 2017; L. J. Core and Lis 2008; L. Core and Adelman 2019). Pol II pause release is associated with the dynamic changes in gene expression that occur during development and differentiation (Boettiger and Levine 2009; Lagha et al. 2013; Saunders et al. 2013; Williams et al. 2015; Marks et al. 2012; Amleh et al. 2009; Abuhashem et al. 2022; Golob et al. 2011). Moreover, promoter-proximal pausing is enriched at genes involved in signaling pathways, including cell cycle, development, and differentiation (Zeitlinger et al. 2007; Tee et al. 2014; Adelman and Lis 2012; Gala et al. 2022), and has emerged as an important link between cell signaling and transcriptional regulation during differentiation (Min et al. 2011; Tastemel et al. 2017; Bai et al. 2013; Elagib et al. 2008). These observations, predominantly in *Drosophila*, zebrafish, or mice, highlight the importance of Pol II pausing in differentiation.

Several factors converge to pause Pol II and mediate its release, including DRB sensitivity-inducing factor (DSIF), which is composed of SPT4 and SPT5. SPT5 is an evolutionarily conserved and essential elongation factor (Wada et al. 1998; Decker 2021; Dollinger and Gilmour 2021; Hartzog et al. 1998) with key roles in maintaining enhancer landscapes, termination, productive elongation, and stabilizing paused Pol II (Hu et al. 2021; Fitz et al. 2020; Cortazar et al. 2019; Parua et al. 2018; Fitz, Neumann, and Pavri 2018; Shetty et al. 2017; Aoi et al. 2021). Despite our understanding of the factors controlling Pol II pausing, examining the role of pausing in the context of differentiation has been challenging because mutating or depleting the factors that regulate pausing (including SPT5) often leads to cell death (Fitz, Neumann, and Pavri 2018; Bai et al. 2013; Rahl et al. 2010; Guo et al. 2000; Tastemel et al. 2017). Thus, it has not been possible to follow cellular differentiation from stem cell to mature cell when pausing is disrupted. Because different sets of genes are paused in various cell states (Min et al. 2011; Adelman and Lis 2012), the specific function of Pol II pausing as cells transition through multiple cell states over the course of differentiation remains unclear.

One well-studied paradigm for differentiation is erythropoiesis, in which hematopoietic stem and progenitor cells (HSPCs) differentiate through a continuum of cell states, beginning with differentiation into lineage-committed erythroid progenitors and ending in terminal differentiation and the production of red blood cells (RBCs) (Orkin and Zon 2008; Cantor and Orkin 2002; Tusi et al. 2018; Zivot et al. 2018; Granick and Levere 1964; Gifford et al. 2006). Adult RBCs are highly specialized cells consisting of ∼95% adult hemoglobin, a complex made up of two β-globin (HBB) and two α-globin (HBA1/2) chains (Weed, Reed, and Berg 1963). Accordingly, *HBB* and *HBA1/2* are highly transcribed during RBC differentiation. Because RBCs expel their nuclei during terminal differentiation, preventing any subsequent transcription, the high level of globin transcription must occur within a tight time window prior to enucleation. Thus, the balanced production of β-globin and α-globin requires precise temporal regulation of the transcriptional programs necessary for effective erythropoiesis. Connections between Pol II pausing and effective erythropoiesis have been observed in multiple settings (Ransom et al. 2004; Bai et al. 2010, 2013; Sawado et al. 2003; Yang et al. 2016; Murphy et al. 2021), but interpretations of these data were constrained due to the lethal effects of mutations that perturb pausing.

We identified a unique opportunity to bypass existing limitations in the study of Pol II pausing through insights we gained from our human genetic studies. We identified loss-of-function mutations in *SUPT5H*, which encodes SPT5, in eight different families with a β-thalassemia trait-like phenotype in the absence of *HBB* mutations. Given that SPT5 regulates Pol II pausing, these mutations presented a unique opportunity to probe the specific function of Pol II pausing during differentiation. Mimicking the pathogenic mutations in human HSPCs undergoing erythroid differentiation, or acutely treating such cells with transcription inhibitors, recapitulated the β-thalassemia phenotype, characterized by reduced levels of β-globin (*HBB*) synthesis relative to alpha-globin (*HBA1/2*) production. During erythropoiesis in healthy human cells, we observed dynamic patterns of Pol II pausing. At the transition from progenitors to precursors, genes involved in the cell cycle were highly paused. In *SUPT5H*-edited cells, we found that Pol II pause release was globally disrupted, and that as cells transitioned from progenitors to precursors, both cell cycle kinetics and the onset of erythroid gene expression programs were delayed. Despite this delay in differentiation, cells effectively underwent terminal differentiation, and cell cycle phase distributions normalized. Together, our results demonstrate that effective erythropoiesis requires both robust transcriptional regulation to maintain globin balance, and Pol II pause release to temporally coordinate the cell cycle and differentiation at a critical transition during erythropoiesis.

## Results

### *SUPT5H* variants associate with an unlinked β-thalassemia phenotype

In the course of searching for the underlying genetic basis of unusual familial blood disorders, we encountered a large pedigree from Italy in which multiple individuals had features of β-thalassemia trait, including an elevated hemoglobin A2 level and signs of β/α-globin chain imbalance, without any mutations in *HBB* identified (unlinked β-thalassemia), despite extensive analyses (Table S1). Whole-exome sequencing across 15 affected and unaffected individuals in this family revealed rare (<0.01% allele frequency in ExAC and GnomAD) loss-of-function mutations in one copy of *SUPT5H*, which encodes the transcription elongation factor SPT5.

Linkage analysis across this family showed a logarithm of the odds score of > 3.5. Further analysis of seven additional pedigrees involving trios with one or two affected individuals with similar phenotypes of unlinked β-thalassemia revealed that all of these families also had loss-of-function *SUPT5H* variants, including nonsense and frameshift mutations. We also identified a rare *SUPT5H* missense variant in this cohort. The mutations were distributed across the entire protein and did not cluster in any particular region (Table S1). Taken together, the *SUPT5H* loss-of-function patient mutations result in a β-thalassemia trait-like phenotype unlinked to *HBB*. While we were conducting these studies, independent pedigrees with similar phenotypes were reported, lending further support to our genetic findings (Charnay et al. 2022; Achour et al. 2020; Lou et al. 2022). Notably, patients harboring these mutations are relatively healthy, exhibiting subtle but consistent phenotypes: essentially no anemia, but clear signs of β/α-globin chain imbalance. Because erythropoiesis is a well-established system, and SPT5 regulates Pol II pausing, these tolerable *SUPT5H* mutations presented an ideal scenario to study the mechanisms by which Pol II pausing regulates differentiation and how its perturbation can cause human disease.

### Pol II pausing is dynamic during human erythropoiesis

Because SPT5 regulates Pol II promoter-proximal pausing, we first assessed Pol II pausing in healthy human HSPCs undergoing erythroid differentiation. For this purpose, we performed native elongating transcript sequencing (NET-seq), which maps Pol II position genome-wide with nucleotide resolution (Mayer et al. 2015), on each day of a 12 day semi-synchronous culture that faithfully recapitulates the major features of *in vivo* erythropoiesis (Figure 1A, 1B, S1A-D) (Giani et al. 2016; Nandakumar et al. 2019; Cantor and Orkin 2002; Perry and Soreq 2002). Nascent gene expression profiles for two healthy donors were highly correlated (Figure S1E), where much of the variability between biological replicates could be attributed to the differentiation trajectory (Figure 1B), and expression analysis was consistent with previous studies (Figure S1F-H) (Ludwig et al. 2019; Georgolopoulos et al. 2021; An et al. 2014).

**Figure 1.**
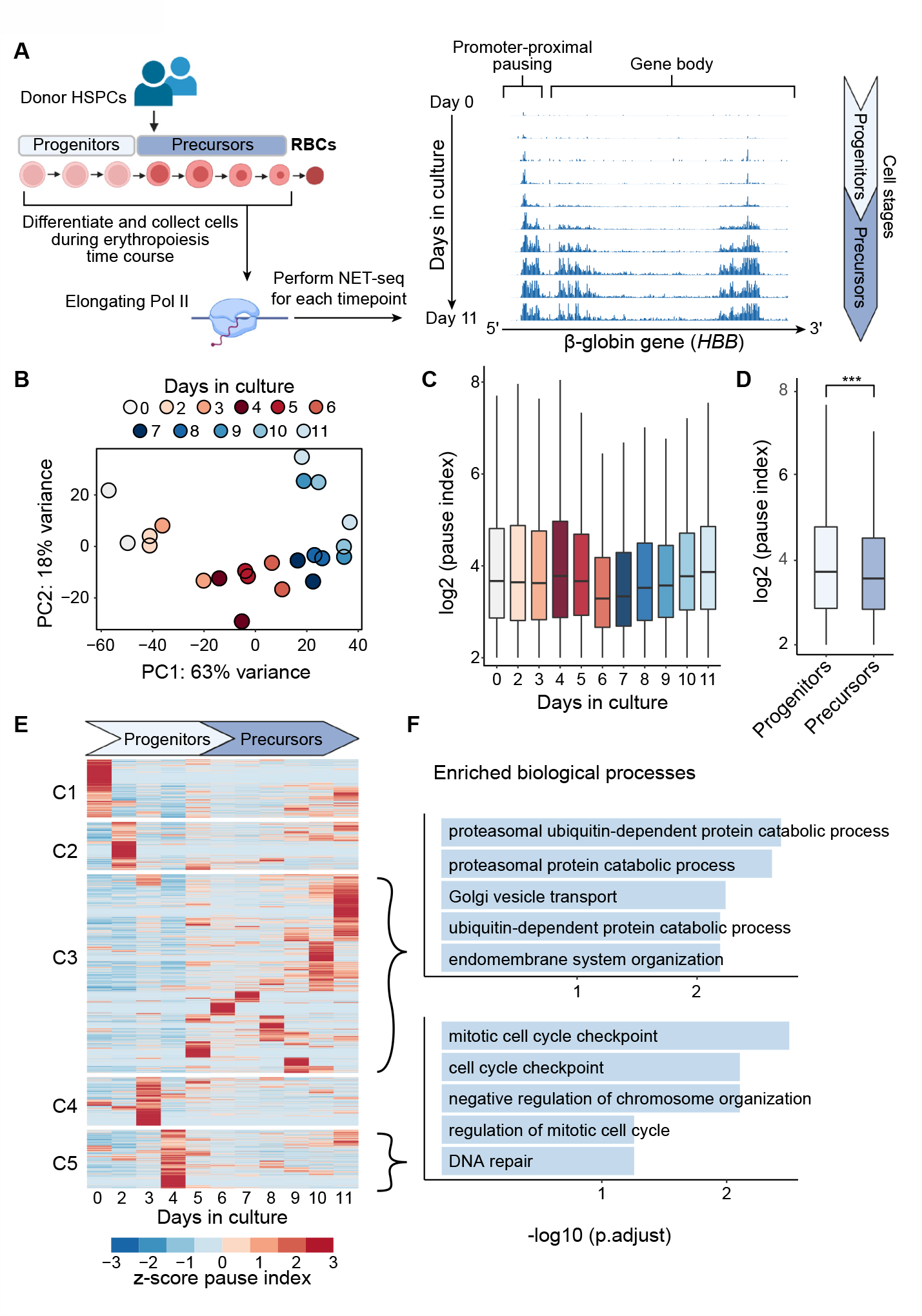
Pol II promoter-proximal pausing is enriched at cell cycle genes during human erythroid differentiation. (A) Schematic of NET-seq time course with daily sampling during human erythropoiesis (left). Transcript per million (TPM) normalized read counts for the *HBB* gene throughout erythropoiesis (right); y axis showing range of 0–15 normalized counts. (B) Principal component analysis plot of NET-seq expression data colored by day of erythroid differentiation; n = 2 donors. (C) Dynamic pausing during erythropoiesis, showing protein coding genes with PI > 4 (n > 3500 genes for all days), significant differences in pausing seen for each day relative to the previous day, except for days 2 and 9; p-values < 0.01 from t-test. (D) Pausing indices of genes in Figure 1C separated by ∼progenitor cells (days 0 to 5, n = 39,466 genes) and ∼precursor cells (days 6 to 11, n = 27,672 genes); t-test. (E) Heatmap of paused genes during erythropoiesis; unsupervised k-means clustering yielded five clusters. Values are z-score of per day average pause index from two donors. (F) Enriched Gene Ontology biological processes for clusters 3 and 5 of paused genes shown in E, other clusters had no significantly enriched biological processes. The top five enriched processes are shown with their - log_10_(p.adjust). P-values: *: p<0.05; **: p<0.01; ***: p<0.001

To investigate Pol II pausing during differentiation, we calculated the pausing indices, the ratio of Pol II promoter region density to Pol II gene body density, for all Pol II protein-coding genes (Figure 1C, Table S2). This metric captures the peak of Pol II density, which represents paused polymerases, and compares it to the overall transcriptional level in the gene body. We identified genes that exhibit a strong promoter bias of Pol II occupancy over the course of differentiation (i.e., pausing index greater than four). Notably, more genes had a high level of promoter-paused Pol II in progenitor cells than in more differentiated precursor cells (Figure 1D) and pausing was highly dynamic as cells transitioned from early progenitors to late-stage erythroblasts (Figures 1C, 1E).

To determine the biological processes governed by the genes that exhibited Pol II pausing during erythroid differentiation, we performed unsupervised k-means clustering followed by GO analysis on the resultant clusters (Figure 1E-F). Notably, cluster 5 genes, which exhibited strong Pol II pausing during the transition from progenitors to precursors (primarily day 4) (Figure 1C, 1E), were significantly enriched for cell cycle processes (Figure 1F). These results suggest that Pol II pausing may play a role in regulating cell cycle progression during erythroid differentiation as cells transition from progenitors into more mature erythroid precursors.

### Editing *SUPT5H* in human HSPCs recapitulates the β-thalassemia phenotype

To analyze how *SUPT5H* loss-of-function mutations impact pausing, we used CRISPR/Cas9 genome editing to perturb *SUPT5H* in human HSPCs from healthy adult donors (Figure 2A) (Shen et al. 2021). To study the effect of this perturbation, edited HSPCs were either (1) plated at low density to form erythroid colonies, such that each colony is derived from a single distinct edited HSPC to obtain a truly heterozygous *SUPT5H* population (Shen et al. 2021); or (2) cultured in bulk using an established differentiation protocol (Giani et al. 2016; Nandakumar et al. 2019). In both cases, we observed predominantly monoallelic editing of *SUPT5H* (Figure 2B, S2A) (Bak et al. 2017). *SUPT5H*-edited cells had reduced SPT5 protein levels (∼50%) relative to control cells in which we targeted *AAVS1*, a genomic region where editing has no impact on hematopoiesis (Figure 2C-D) (Cromer et al. 2022). Acute SPT5 depletion in cell lines co-occurs with a reduction in Pol II levels (Hu et al. 2021; Aoi et al. 2021; Fong et al. 2022). Similarly, *SUPT5H*-edited cells had modestly lower Pol II levels (Figure S2B). Globin expression was decreased and the ratio of *HBB* to *HBA* mRNA was lower in *SUPT5H*-edited cells than in the control cells (Figure 2E, S2C-D, Table S3). This globin imbalance recapitulates the major features of the β-thalassemia phenotype observed in individuals carrying *SUPT5H* mutations, enabling us to study the mechanistic basis of this phenotype *in vitro*.

**Figure 2.**
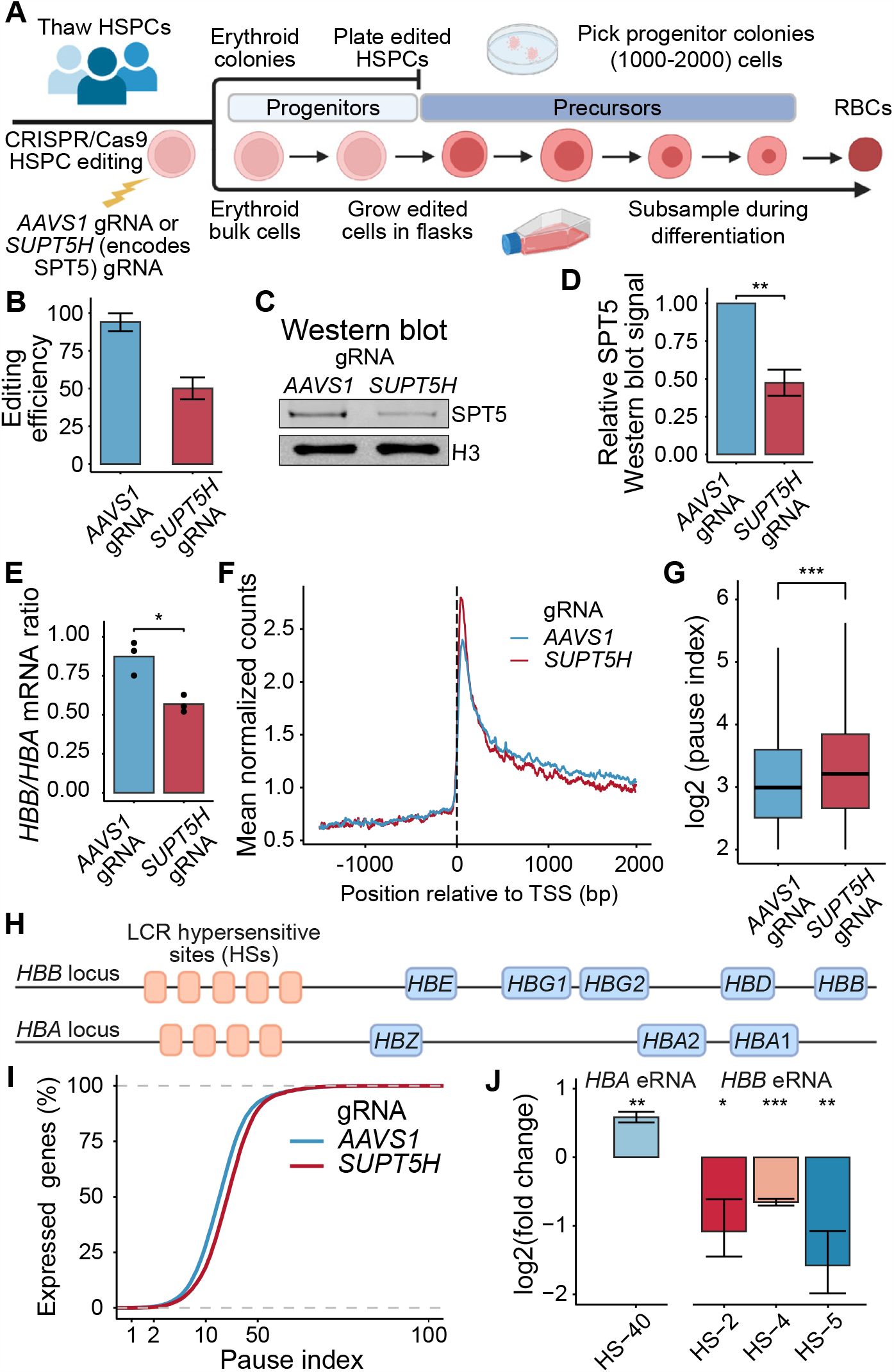
Pol II promoter-proximal pause release is hindered in human hematopoietic cells upon perturbation of *SUPT5H*. (A) Schematic of genome editing of human HSPCs from healthy donors in an isogenic setting. Edited cells were either plated at low density to form erythroid colonies (erythroid clonal cell population) or cultured in bulk. (B) Editing efficiency of gRNAs evaluated by CRISPR amplicon sequencing in bulk HSPCs from three donors. Data are represented as means ± SEM. (C) Western blot of HSPCs that were treated with gRNAs targeting the *SUPT5H* gene or an *AAVS1* control. H3 serves as a loading control. (D) Western blot quantification of SPT5 knockdown. Signal was normalized against H3 and *AAVS1*. Data are represented as means ± SEM; n=3 donors, t-test. (E) Ratio of HBB mRNA relative to HBA mRNA for erythroid colonies; n =3 independent experiments for each gRNA, t-test. (F) Average normalized sense-strand NET-seq signal around transcription start site (TSS) for Pol II protein-coding genes comparing *AAVS1* control and *SUPT5H*-edited cells; n=3 donors. (G) Comparison of pausing indices between *AAVS1* control and *SUPT5H*-edited cells reveals significantly higher pausing in *SUPT5H*-edited cells; n=3 donors, t-test. (H) Schematic of the *HBB* and *HBA* loci including the *HBB* and *HBA* locus control regions (LCR) (i.e enhancer regions). (I) Cumulative distribution of the pausing indices of Pol II protein-coding genes; p < 0.0001 from a two-sample Kolmogorov-Smirnov test. (J) Enhancer normalized read counts in hypersensitive (HS) sites of the HBB and HBA LCR. The other HS sites do not show significant differences between *AAVS1* control and *SUPT5H*-edited cells (Figure S2H-I); n=3 donors, t-test. P-values: *: p<0.05; **: p<0.01; ***: p<0.001

### Editing *SUPT5H* globally disrupts Pol II pause release

To determine the transcriptional effects of perturbing *SUPT5H*, we performed NET-seq on *SUPT5H*-edited cells on day 8 of differentiation (Figure S2E), a time point when globin expression levels are rapidly increasing, and the erythroid cultures have expanded such that cell number is not limiting (Figure S1H). Because SPT5 is a regulator of Pol II pausing, we examined the normalized NET-seq signal around promoter regions. In *SUPT5H*-edited cells, we observed an enrichment of Pol II occupancy at promoter-proximal regions, along with a reduction in downstream genic regions, relative to control cells (Figure 2F). *SUPT5H*-edited cells had a greater percentage of genes with a high level of promoter-proximal pausing (Figure 2G, 2I, Table S4), suggesting that Pol II pause release into productive elongation was hindered. Consistent with our analysis of mature mRNA, the ratio of *HBB* NET-seq signal to *HBA* signal was significantly reduced (Figure S2F). Examination of the NET-seq signal around globin promoter regions revealed no significant changes in pausing at these loci relative to control cells (Figure S2G). Next, we examined non-coding enhancer-derived RNAs (eRNAs) in the enhancer regions (LCR) of *HBB* and *HBA* (Figure 2H). The levels of eRNAs within the *HBA* LCR were elevated in *SUPT5H*-edited cells (Figure 2J), whereas the levels of eRNAs within the *HBB* LCR were significantly reduced (Figure 2J, S2H-I). Given that eRNA transcription is a reliable marker of active enhancers (Lam et al. 2014; Kowalczyk et al. 2012), our results suggest that *HBB* LCR activity is reduced when SPT5 is perturbed, potentially contributing to the reduced *HBB* expression observed in patients.

### SPT5 regulates the temporal expression of erythroid differentiation genes

In *SUPT5H*-edited cells, Pol II pause release was disrupted across many genes, suggesting that the effects of perturbing *SUPT5H* are not limited to globins and may affect gene expression on a global level. RNA-seq analysis of differentiated cells from erythroid colonies (Figure 3A, S3A, S3D) revealed that downregulated genes were most enriched for GO terms related to the cell cycle and erythroid differentiation (Figure 3B). Surprisingly, RNA-seq analysis of cultured *SUPT5H*-edited cells during *in vitro* erythropoiesis (days 2 to 10 in culture) (Figure 3C, S3B) revealed very few differentially expressed genes (DEGs) on most days, with the striking exception on day 4 at the transition from progenitors to precursors, when thousands of DEGs were detected (Figure 3C-D). Consistent with the colony results (Figure 3A-B), genes downregulated on day 4 in erythroid cultures were enriched for cell cycle and erythropoiesis genes, including CDK1, a major regulator of cell cycle transcriptional programs (Enserink and Kolodner 2010) and master regulators of human erythropoiesis, KLF1 and GATA1 (Perkins et al. 2016; Suzuki, Shimizu, and Yamamoto 2011) (Figure 3D, S3C). Because cell cycle genes exhibited stage-specific pausing at this same time point during erythropoiesis (Figure 1E-F), we asked whether the changes in gene expression could be due to hindered pause release upon perturbation of *SUPT5H*. Indeed, genes with reduced expression on day 4 significantly overlapped with genes with strong stage-specific Pol II pausing in unedited cells (p = 5 × 10^−5^, Fisher’s exact test) (Figure 1E), whereas genes with increased expression on day 4 showed no significant overlap. Additionally, pausing indices in unedited cells, particularly on day 4, were significantly higher for genes with reduced expression upon *SUPT5H* perturbation than for genes with increased expression (Figure 3E). Taken together, our results suggest that the gene expression programs governing the transition from progenitors to erythroid precursors are regulated by pausing.

**Figure 3.**
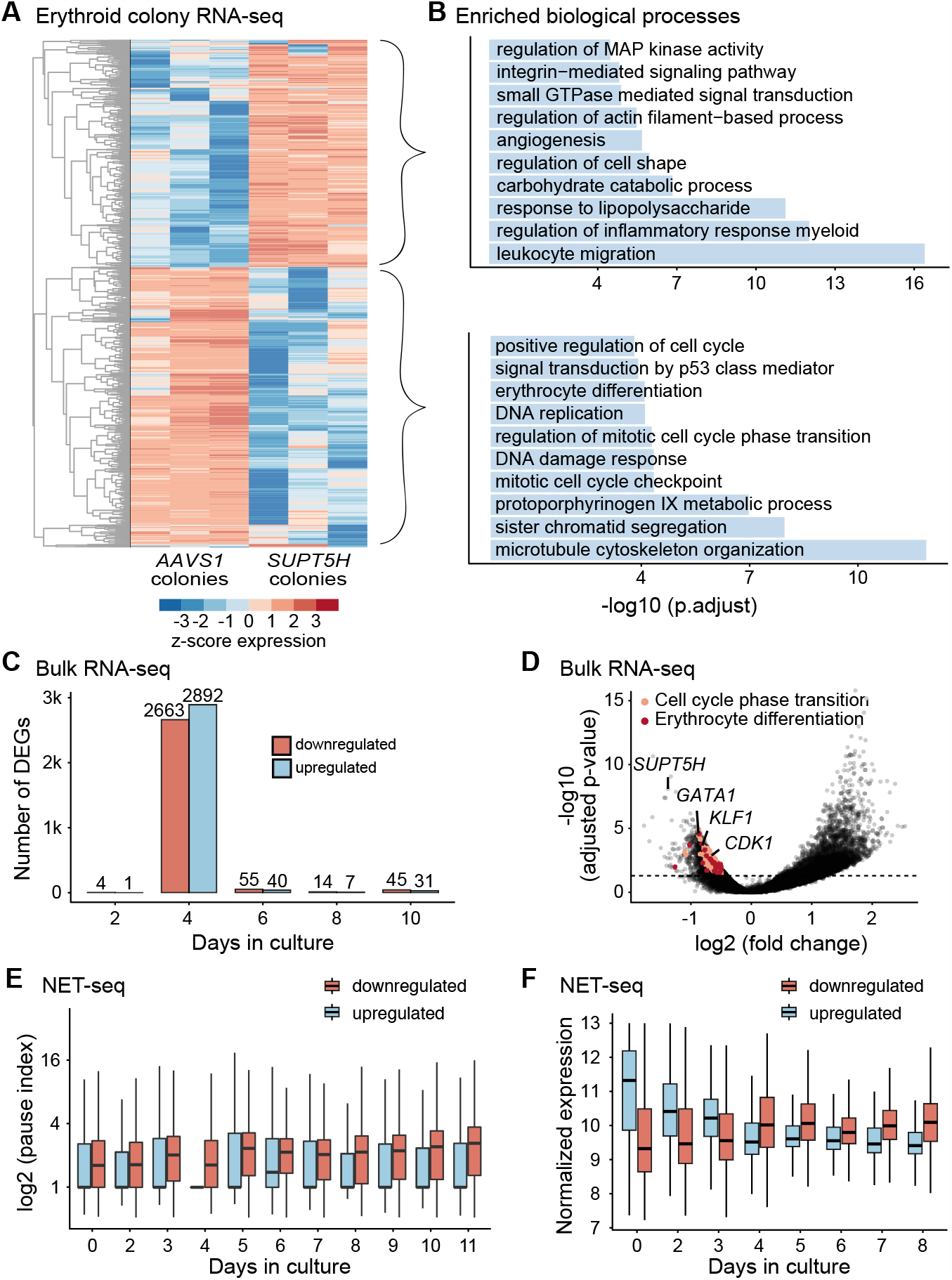
Perturbing *SUPT5H* delays erythroid gene expression programs. (A) Heatmap of differentially expressed genes (n= 1,750, adjusted p-value <0.05) from erythroid colonies from three independent experiments on single colonies comparing control and *SUPT5H*-edited cells. (B) Gene Ontology biological processes for differentially expressed genes (upregulated and downregulated) of erythroid colonies (comparing *AAVS1* and *SUPT5H*-edited cells). The top ten enriched representative processes are shown with their -log_10_(p.adjust). (C) Number of differentially expressed genes from RNA-seq on bulk edited erythroid culture cells comparing *AAVS1* and *SUPT5H*; days 2, 4, 6, 8, and 10 of erythroid differentiation, colored by increased or decreased expression, n=3 donors, adjusted p-value < 0.05. (D) Volcano plot of erythroid culture cells showing decreased expression for genes including *SUPT5H, GATA1, KLF1*, and *CDK1* in *SUPT5H*-edited cells on day 4 of erythroid differentiation. Horizontal line falls at adjusted p-value 0.05. (E) Comparison of NET-seq pausing indices in wild-type cells (from time course, Figure 1), categorized by differentially expressed genes (upregulated, n=2892 or downregulated, n=2663) in *SUPT5H*-edited cells on day 4 of erythroid differentiation. Distributions are significantly different for all days except day 6, t-test, p < 0.01. (F) NET-seq normalized expression z-scores from wild-type cells (from time course, Figure 1), categorized by differentially expressed genes (upregulated or downregulated) in *SUPT5H*-edited cells on day 4 of erythroid differentiation.

We next asked how *SUPT5H* editing would alter the dynamic gene expression changes that occur as cells transition from early erythroid progenitors to more terminally differentiated erythroid populations, when globins and other erythroid-specific genes are rapidly induced (Tusi et al. 2018; Nestorowa et al. 2016). Genes with reduced expression on day 4 in *SUPT5H*-edited cells were more highly expressed on or after the transition from progenitors to precursors in unedited cells. By contrast, genes whose expression increased on day 4 were more highly expressed prior to the transition to precursors in unedited cells (Figure 3F). Thus, gene expression programs in progenitor cells were expressed for longer in *SUPT5H*-edited cells, whereas the onset of terminal erythroid gene expression programs were delayed. Notably, the delay was transient, with very few DEGs detected before or after this time point (Figure 3C), suggesting that the kinetics of differentiation were altered but that *SUPT5H*-edited cells recovered as differentiation progressed. Thus, SPT5 controls the timing of the induction of cell cycle and differentiation gene expression programs, linking Pol II pause release to the efficient transition from erythroid progenitors to precursors.

### SPT5 promotes the transition from progenitors to erythroid precursors

Based on the altered gene expression in differentiating *SUPT5H* edited cells, we hypothesized that differentiation timing was affected in these cells. To test this, we measured canonical differentiation markers using flow cytometry (Figure 4A-B). Human HSPCs express CD34, which is lost once cells enter a late progenitor stage. Erythroid progenitors acquire expression of CD36 and CD71, which in more differentiated erythroid precursors are co-expressed with glycophorin A (GYPA; also known as CD235a). During the late stages of terminal maturation expression of CD36 and CD71 are gradually downregulated (Mao et al. 2016; Neildez-Nguyen et al. 2002; Fajtova et al. 2013; Okumura, Tsuji, and Nakahata 1992; J. Li et al. 2014; Yan et al. 2021). Consistent with our hypothesis, *SUPT5H*-edited samples retained a significant population of CD34+ progenitors as cells transitioned from progenitors to precursors, demonstrating that *SUPT5H*-edited cells retained a more immature progenitor population than control cells (Figure 4C-D). At the same time point, the proportion of cells transitioning from progenitors to erythroid precursors (CD34+/CD36+) decreased. By contrast, at other time points during erythropoiesis we observed no significant differences in the expression of these markers between *SUPT5H*-edited and control cells. Thus, perturbing SPT5 levels transiently delays differentiation as erythroid progenitors transition to precursors.

**Figure 4.**
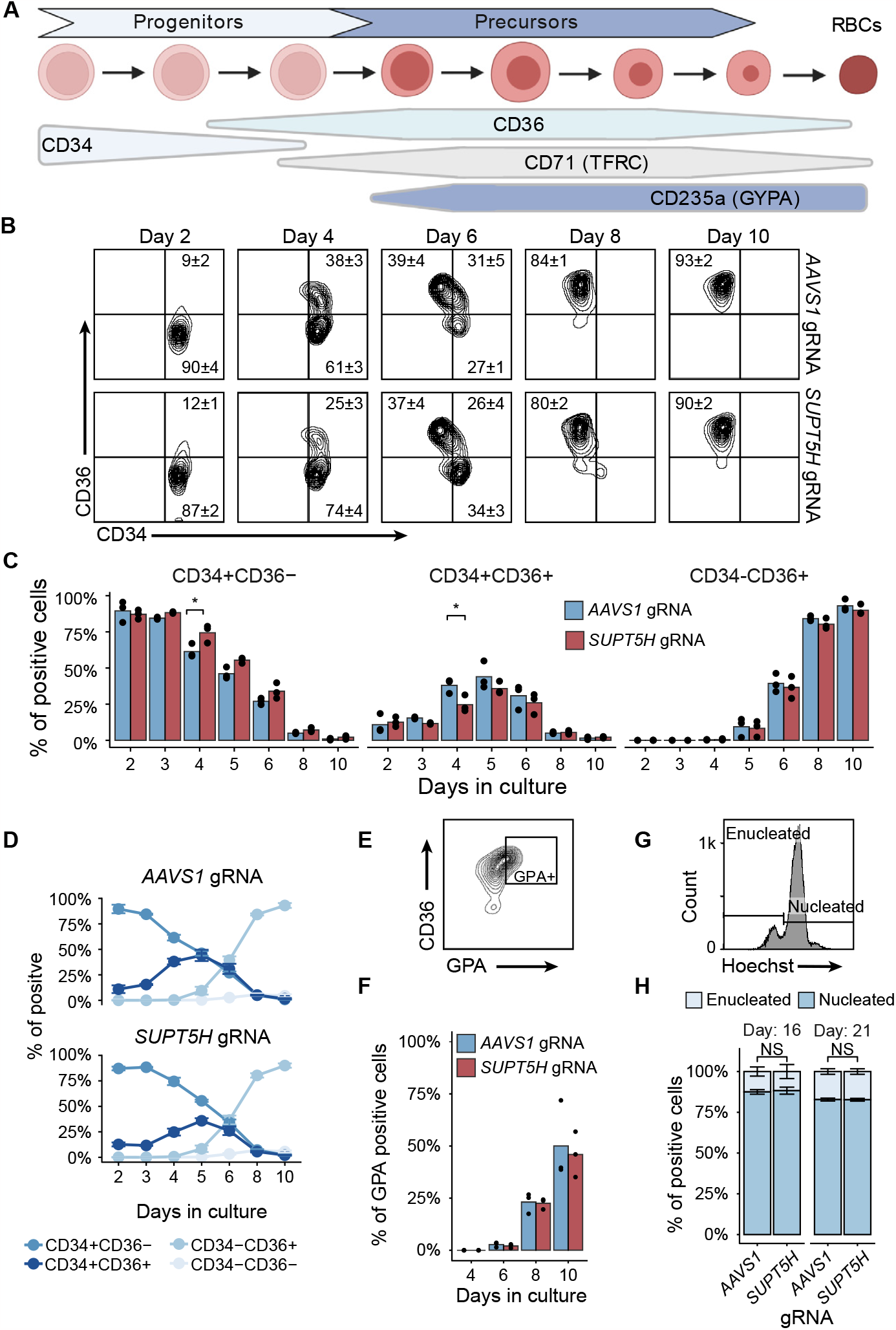
Reduced levels of *SUPT5H* result in a stage-specific delay in erythroid differentiation. (A) Schematic outline of canonical differentiation markers during *ex vivo* erythroid differentiation of human HSPCs to terminal differentiation, just prior to enucleation. (B) The maturation of erythroid precursor cells from adult CD34+ HSPCs was assessed with FACS by serially tracing the expression of erythroid lineage markers CD34 and CD36, comparing *SUPT5H*-edited cells and the *AAVS1*-edited control cells. Days 2, 4, 6, 8, and 10 of erythroid differentiation in culture; n = 3 donors, showing means ± SEM. (C) Quantification of cell populations transitioning from less mature (CD34+/CD36-) to a more mature cell population (CD34+/CD36+ followed by CD34-/CD36+), comparing *SUPT5H*-edited cells and the *AAVS1*-edited control cells; t-test, p < 0.05. (D) Line plot showing the trajectory of erythroid differentiation as cells transition from progenitors to precursors. Trajectory of *SUPT5H*-edited cells is perturbed relative to *AAVS1*-edited control cells, especially on days 4 and 5. (E) Representative FACS plot of erythroid terminal differentiation markers CD36 and GPA of *SUPT5H*-edited cells after 21 days of differentiation. (F) Quantification of acquisition of GPA during erythroid terminal differentiation for *SUPT5H*-edited cells and the *AAVS1*-edited control cells. NS: not significant (t-test). (G) Representative FACS plot of erythroid enucleation using Hoescht stain to measure nuclear content; plot shows *SUPT5H*-edited cells on day 21 of erythroid terminal differentiation. (H) Quantification of enucleation during erythroid terminal differentiation showing no significant difference (t-test) between *SUPT5H*-edited cells and the *AAVS1*-edited control cells. Measured on days 16 and 21; data are represented as means ± SEM.

We next asked whether *SUPT5H*-edited cells exhibit any differences during terminal erythroid maturation. The levels of several canonical erythroid terminal maturation markers (GYPA, CD36, and CD71) did not differ between *SUPT5H*-edited and control cells (Figure 4E-F, S4A-D). Similarly, we observed no difference between edited and control cells in the timing or the extent to which the cells enucleated (Figure 4G-H) or in the morphology of terminal erythrocytes (Figure S4E). Thus, cells experiencing a transiently delayed transition from progenitors to erythroid precursors recovered and effectively underwent terminal differentiation, consistent with the absence of anemia in individuals with *SUPT5H* loss-of-function mutations.

### Perturbing *SUPT5H* causes stage-specific changes in the cell cycle

The downregulation of genes involved in the cell cycle in *SUPT5H*-edited cells led us to ask whether the cell cycle is altered in *SUPT5H*-edited cells during the transition from progenitors to precursors. Consistent with previous studies describing cell cycle remodeling during murine erythropoiesis (Socolovsky 2022; Hwang et al. 2017), we observed a rapid increase in the S-phase cell population, and a reciprocal decrease in the G0/G1 population, in control cells (Figure 5A-B). By contrast, in *SUPT5H*-edited cells, we observed a prolonged shift in cell cycle dynamics with a higher percentage of cells in G0/G1 (58±2% vs 45±3%) and a lower percentage in S (39±1% vs 52±2%) relative to control cells (Figure 5A-C, S5D). These changes were especially pronounced on day 4 in *SUPT5H*-edited cells, in which cell cycle genes were downregulated simultaneously with the delay in differentiation (Figure 4C-D). To broadly confirm the observed changes in cell cycle remodeling, we measured proliferation via luminescence (Figure S5A), and found proliferation significantly increased as cells transitioned from progenitors to precursors in *SUPT5H*-edited cells (Figure 5D, S5B-C). The difference in proliferation dissipated as cells progressed to the precursor stage, likely due to the rapid shift in cell cycle dynamics where control cells enter a highly proliferative state sooner than *SUPT5H*-edited cells (Figure 5B, S5A). Furthermore, on day 4, S phase was significantly faster in *SUPT5H*-edited cells than in control cells (Figure 5E-F). Thus, *SUPT5H*-edited cells experience a pronounced disruption to their cell cycle at the transition from progenitors to precursors, mirroring the delay in differentiation kinetics.

**Figure 5.**
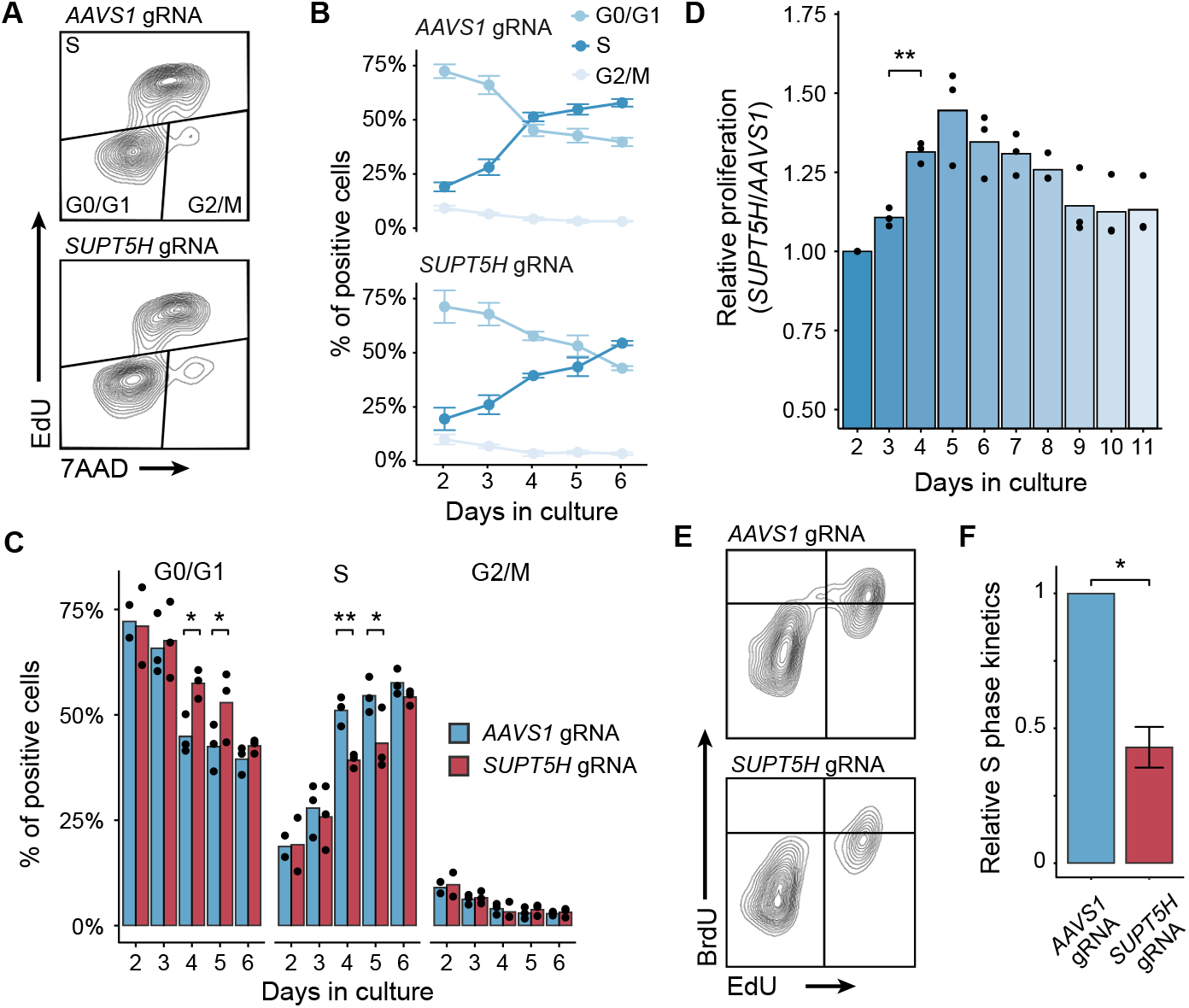
Perturbing *SUPT5H* leads to stage-specific delay in the cell cycle programs. (A) Representative FACS plot of EdU signal and DNA staining (7AAD) to measure cell cycle populations. Plot shows day 4 of the *SUPT5H*-edited cells and the *AAVS1*-edited control cells. FACS plots for all sampled days are shown in Figure S5D. (B) Line plot of the switch in cell cycle programs during *ex vivo* erythroid differentiation as human erythroid progenitors transition to precursors for *SUPT5H*-edited and *AAVS1*-edited cells. Data are from FACS analysis of EdU signal and DNA staining (7AAD) (Figure S5D). (C) Quantification of cell cycle populations during the erythroid progenitor stages in *SUPT5H*-edited cells relative to *AAVS1*-edited control cells; t-test. (D) Relative cell proliferation in *SUPT5H*-edited cells measured by ATP bioluminescence during *ex vivo* erythroid differentiation going from human erythroid progenitors to terminal differentiation. *SUPT5H*-edited cell signal relative to *AAVS1*-edited control cells, days 3 to 11 of erythroid differentiation in culture. Nucleofection of gRNA occurred on day 1 in culture and values are normalized against day 2 (24 hrs post-nucleofection). (E) Representative FACS plot of BrdU and EdU dual labeling to measure cell cycle kinetics. Plot shows day 4 of the *SUPT5H*-edited cells and *AAVS1*-edited control cells. (F) Quantification of cell cycle kinetics on day 4 of erythroid differentiation shows a significant decrease in S-phase kinetics in *SUPT5H*-edited cells compared to the *AAVS1*-edited control cells; S phase length (Ts) = Ti/(P_L_/P_S_), where Ti = EdU labeling time, Ps = EdU+/BrdU+ cell population, and P_L_ = EdU+/BrdU-cell population (Martynoga et al. 2005); data are represented as means ± SEM, t-test. P-values: *: p<0.05; **: p<0.01; ***: p<0.001

### Inhibiting transcription is sufficient to recapitulate the β-thalassemia phenotype

Our observations up to this point demonstrated that editing *SUPT5H* disrupts erythropoiesis and globin expression in multiple ways: hindering pause release into productive elongation, reducing *HBB* LCR activity, transiently delaying differentiation, and altering cell cycle dynamics at specific stages of erythropoiesis. Therefore, we next sought to determine which of these changes contribute to the patient β-thalassemia phenotype of globin imbalance. To decouple the delays in differentiation and the cell cycle during the progenitor to precursor transition (day 4) from the direct effects of reduced SPT5 during the period of high globin expression (day 8+) (Figure S1H), we used SPT5-Pol II small molecule inhibitors (SPIs) to inhibit SPT5 function on day 8 during the differentiation of unedited cells (Bahat et al. 2019). Modest SPI levels were used to reflect the ∼0.5 fold SPT5 depletion in *SUPT5H*-edited cells (Figure S6A-B). Moreover, we asked whether the general disruption of pause release or transcription initiation on day 8 would similarly result in globin imbalance. We analyzed the effects of three types of small molecule inhibitors: SPIs, inhibitors of Pol II pause release (flavopiridol and DRB), and an inhibitor of transcription initiation (triptolide) (Figure 6A, 6F, S6C-D) on day 8 of differentiation. NET-seq analysis revealed the expected transcriptional phenotypes of these inhibitors (Figure 6B-D, 6G, S6E). Treatment with both SPIs recapitulated the results seen in *SUPT5H*-edited cells of increased Pol II promoter-proximal pausing (Figure 6G, S6E, Table S5). Pause release inhibitors also lead to increased Pol II promoter-proximal pausing and the transcription initiation inhibitor led to decreased Pol II occupancy in both promoters and gene bodies (Figure 6B-D, Table S6). Strikingly, analysis of *HBB*/*HBA* ratios revealed a globin imbalance in all perturbations (Figure 6H-I, S6F-H, Table S7-8). Even very short (15 min) treatments with DRB or triptolide decreased *HBB*/*HBA* mRNA ratios, revealing how sensitive globin transcription is to transcriptional perturbations (Figure 6E). Overall, these results indicate that disrupting transcription is sufficient to produce a globin imbalance, suggesting that the transient cell cycle and differentiation effects in *SUPT5H*-edited cells may not contribute to the globin imbalance observed in the patient β-thalassemia phenotype and these effects earlier in differentiation are likely compensated for during later stages of erythropoiesis.

**Figure 6.**
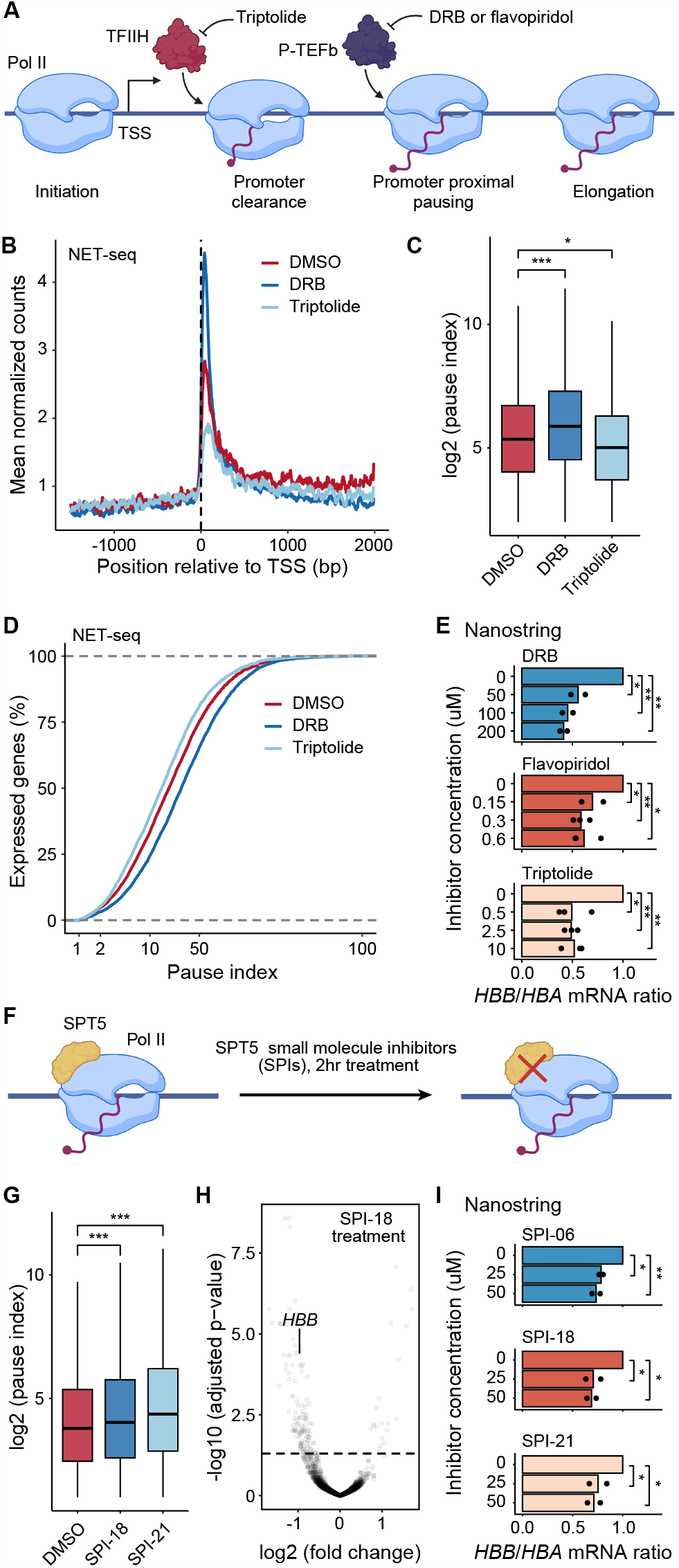
Transcription inhibitors disrupt globin balance and recapitulate the β-thalassemia phenotype. (A) Schematic outline showing the stages of transcription that are impacted by different transcription inhibitors. (B) Average sense-strand normalized NET-seq reads around transcription start site (TSS) for expressed Pol II protein-coding genes comparing DMSO control and transcription inhibitors, DRB and triptolide; cells treated on day 8 of erythroid differentiation, n = 2 donors. (C) Comparison of pausing indices between DMSO control and transcription inhibitors, DRB and triptolide; t-test. (D) Cumulative distribution of the pausing indices of Pol II protein-coding genes; p < 0.0001 from a two-sample Kolmogorov-Smirnov test. (E) Ratio of *HBB* relative to *HBA* calculated from direct mRNA levels comparing the DMSO control and transcription inhibitor–treated cells; 15-minute treatment, data are represented as the mean, t-test. (F) Schematic of SPT5-Pol II small-molecule inhibitor (SPI) perturbation of Pol II–SPT5 interactions. (G) Comparison of pausing indices between DMSO control and two SPIs (SPI-21 and SPI-18); treated on day 8 of erythroid differentiation, t-test. (H) Volcano plot of SPI-18 treated cells showing decreased *HBB* expression. Cells were treated on day 8 of erythroid differentiation for 2 hours. Horizontal line falls at adjusted p-value of 0.05. (I) Ratio of *HBB* relative to *HBA* calculated from direct mRNA levels comparing the DMSO control and SPI treated cells; 2-hour treatment, data are represented as the mean, t-test. P-values: *: p<0.05; **: p<0.01; ***: p<0.001

## Discussion

In this study, we identified and recapitulated a heterozygous loss-of-function mutation in the *SUPT5H* gene, which encodes the transcription elongation factor SPT5. Patients with mutations in *SUPT5H* exhibited phenotypes consistent with unlinked β-thalassemia. Given that erythropoiesis is a well-established system for studying differentiation, and the heterozygous *SUPT5H* mutation is viable in our *in vitro* studies as well as in living individuals, this study provides new insight into how perturbations to pausing can result in a distinct human phenotype, and how promoter-proximal Pol II pausing regulates gene expression changes during differentiation (Figure 7).

**Figure 7.**
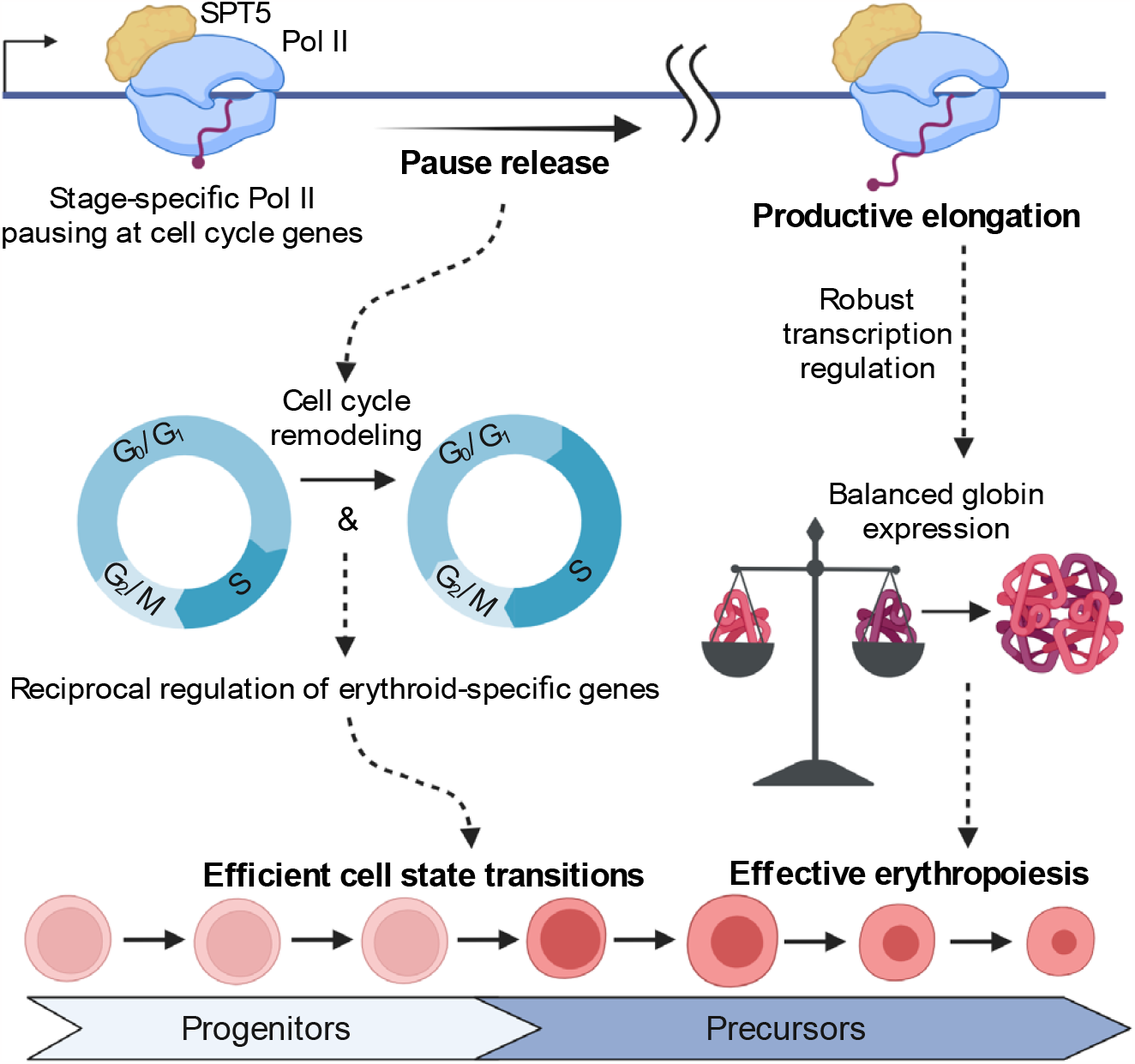
The multiple roles of SPT5 during erythroid differentiation. Pol II pausing coordinates the cell cycle and differentiation, promoting efficient cell state transitions during erythropoiesis. Effective erythropoiesis requires robust transcriptional regulation to ensure balanced globin expression during erythroid differentiation.

By recapitulating the *SUPT5H* loss-of-function mutation observed in humans, we found that hindered pause release resulted in a transient delay from progenitors to erythroid precursors during differentiation (Figure 4B-D). The delay in erythroid differentiation included postponed globin expression (Figure S2D) and other erythroid-specific genes, potentially contributing to the patient phenotype by tightening the window for globin expression prior to enucleation. Nevertheless, *SUPT5H* loss-of-function mutations impact *HBB* expression to a greater extent than *HBA*. We found that *HBB* was more susceptible than *HBA* to transcriptional perturbations (Figure 6E, 6I), particularly at late stages of erythropoiesis when high expression demands may render globins especially vulnerable to such perturbations. Due to the many differences in how *HBB* and *HBA* expression are regulated (Mettananda, Gibbons, and Higgs 2016, 2015), multiple global and local regulatory elements likely impact how SPT5 differentially regulates globin expression. Interestingly, *HBB* expression has been shown to be regulated by the recruitment of SPT5 via the *HBB* LCR (Bender et al. 2012; Song et al. 2010), suggesting that the reduced *HBB* LCR activity we observed might be responsible for the reduced *HBB* expression. Together, our findings provide an example of coordinated gene expression being differentially affected by a general transcription factor, SPT5. This is consistent with previous patient studies showing that mutations in general transcription factors have different effects on *HBA* and *HBB* expression where thalassemia phenotypes were attributed to the perturbed transcription of globin genes (Viprakasit et al. 2001; Theil et al. 2017). Collectively, these observations suggest that further investigation into how general transcription factors regulate gene expression within the context of global and local regulatory elements could provide greater insight into the mechanisms by which genes are differentially expressed.

Finally, our findings have broader implications concerning the role of pausing in proliferation and differentiation. During erythropoiesis, pausing was dynamic (Figure 1C), highlighting the fact that specific genes are paused in various cell states, and consistent with a role for pausing in tuning specific gene expression programs (Gilchrist et al. 2012; Hah et al. 2011; Lin et al. 2011). During differentiation, the genes that exhibited stage-specific pausing were enriched in signal-responsive pathways (Figure 1E-F). For instance, when cells were transitioning from progenitors to precursors, cell cycle genes were highly paused. The transition from progenitors to precursors was marked by a rapid switch in cell cycle and erythroid gene expression programs (Figure 4C-D, 5B), but when pause release was hindered, the timing of this rapid transition was delayed (Figure 4C-D, 5B), suggesting that pausing acts as a temporal regulator, coordinating gene expression programs as cells transition from progenitors to erythroid precursors, and emphasizing that pausing acts as a kinetic bottleneck, determining the rate of cell-state transitions during differentiation (Jonkers and Lis 2015; Abuhashem et al. 2022). These observations tie together numerous observations of the interdependence of differentiation and the cell cycle (Pop et al. 2010; Sankaran et al. 2012; Hojun Li et al. 2019; Tallack et al. 2009) with the enrichment of Pol II pausing at genes involved in the cell cycle, differentiation, and other signal-responsive pathways (Adelman and Lis 2012). In summary, by using human genetics to identify and recapitulate tolerable loss-of-function *SUPT5H* mutations, and following differentiation throughout erythropoiesis, we found that rapid and synchronous transitions in cell cycle and differentiation dynamics are temporally coordinated by pausing. Given that the coordination of proliferation and differentiation is a fundamental process of metazoan multicellularity, and promoter-proximal pausing is a metazoan-specific regulatory step (Peterlin and Price 2006; Dollinger and Gilmour 2021; Chivu et al. 2023), further studies elucidating the connection between pausing and the coordination of proliferation and differentiation could provide insight into the mechanisms that govern cell fate specification.

## Data Availability

All data produced in the present study are available upon reasonable request to the authors.

## Acknowledgements

We thank Brendan Smalec, Andreas Mayer, and Hojun Li for helpful comments and critical reading of the manuscript. We are grateful to the Churchman and Sankaran lab members for engaging discussions and support. Figure schematics were created with BioRender.com. D.J.M. was supported by the National Institute of Health Ruth L. Kirschstein Postdoctoral Fellowship F32-GM125238. H.E.M. was supported by a National Science Foundation Graduate Research Fellowship DGE 1745303. R.I. was supported by EMBO fellowship ALTF 2016-422 and NIH/NIGMS T32 postdoctoral training grant GM007748-44. C.S. was supported by the Charity Action Medical Research grant GN2855. This work was supported by National Institutes of Health grants R01-HG007173 (L.S.C.), R01-DK103794 (V.G.S.), and R01-HL146500 (V.G.S.), the New York Stem Cell Foundation (NYSCF) (V.G.S.), a gift from the Lodish Family to Boston Children’s Hospital (V.G.S.), and the Ellison Foundation (V.G.S). V.G.S. is a NYSCF-Robertson Investigator.

## Author contributions

D.J.M., V.G.S., and L.S.C. conceived and designed the experiments. D.J.M. performed the experiments. D.J.M. and C.F. performed FACS of human CD34+ cells for edited and NET-seq WT data, respectively. D.J.M. and A.C. performed FACS for the fluorescent proliferation staining data. D.J.M. and H.E.M. performed western blots on WT and *SUPT5H*-edited cells, respectively. D.H., C.B., G.G, V.B., M.D.C., C.S., N.R., M.P., N.B.A.R., J.C.U and V.G.S. identified and sequenced β-thalassemia patients with loss-of-function *SUPT5H* mutations. D.J.M., R.I., and H.E.M. optimized spike-in conditions and analysis. D.J.M., H.E.M., R.I. and K.C. performed data analysis. D.J.M., V.G.S., and L.S.C. interpreted the results and wrote the manuscript with input from all authors.

## Methods

### Lead contact

Further information and requests for reagents may be directed to, and will be fulfilled by, the lead contact.

### Materials availability

Unique reagents generated in this study are available from the lead contact without restriction.

### Data and code availability

The WES data will be made available upon publication in the dbGaP database (http://www.ncbi.nlm.nih.gov/gap).

Sequencing data will be made available upon publication in the GEO database (https://www.ncbi.nlm.nih.gov/geo/).

Computational code will be made available on GitHub upon publication (https://github.com/churchmanlab).

## Method Details

### B-thalassemia patient data

The patients described in this paper are part of a rare blood disorder cohort, in several family members thalassemia screening revealed microcytic, hypochromic anemia and elevated HbA 2 consistent with β-thalassemia trait (Table S1). DNA obtained from peripheral blood samples of the patients was subjected to whole exome sequencing (WES) to identify potential pathogenic variants.

### Study approval

All family members provided written informed consent to participate in this study. The study was approved by the Institutional Review Boards of Boston Children’s Hospital. The human study protocols were also approved by the Institutional Review Boards of Boston Children’s Hospital.

### WES and related genetic analyses

A rare blood disorder cohort was studied using WES, which was performed as previously described (Abdulhay et al. 2019; Polfus et al. 2016; Kim et al. 2017; Khajuria et al. 2018; Ulirsch et al. 2018). Peripheral blood samples were obtained from the patients described in this paper and WES was performed using genomic DNA obtained from the blood samples. The resultant variant call file (in hg19 coordinates) was annotated with VEP v89 (McLaren et al. 2016) and rare variants (based on ExAC v0.3.1 and GnomAD r2.0.2; http://gnomad.broadinstitute.org/) (Lek et al. 2016) were identified using a combination of the Genome Analysis Toolkit, Bcftools, and Gemini (McKenna et al. 2010; Heng Li 2011; Paila et al. 2013). No rare (<0.01% allele frequency in ExAC and GnomAD) loss-of-function or missense variants were identified in any transcription factors known to regulate HBB, including those known to cause β-thalassemia (GATA1, GATA2, KLF1, KLF3, ERCC2, CCND3, CTSA, PCIF1, PLTP, MMP9, TNNC2, ZFPM1, NFE2, FOG1, SOX6, SP1, and AKL4). We next expanded our search and noted a likely pathogenic variant in *SUPT5H* harbored by these patients but absent from ExAC and GnomAD (Table S1). The Hb, MCV and MCH in *SUPT5H* carriers is markedly reduced compared to normal family members, while the HbA2 is increased, similar to β-thalassemia trait. All mutations were confirmed from genomic DNA samples of the patients or family members by Sanger sequencing.

### Primary HSPC culture and colony-forming cell assay

The Institutional Review Board at Boston Children’s Hospital approved the use of de-identified human HSPCs. The CD34-enriched HSPCs from healthy human adult donors were obtained from the Fred Hutchinson Hematopoietic Cell Processing and Repository (Seattle, USA). The HSPCs were thawed and cultured in medium using an established differentiation protocol that faithfully recapitulates the major features of *in vivo* erythropoiesis (Giani et al. 2016; Nandakumar et al. 2019). Subsamples were collected from each day of a single continuous culture of human adult HSPCs. For colony-forming cell assays, HSPCs were plated at low density (500 cells/mL) in semi-solid medium (MethoCult Optimum methylcellulose medium, StemCell Technologies, Inc.) to form erythroid BFU-E colonies (as well as other hematopoietic colony types), such that each colony is derived from a single distinct edited HSPC (clonal cell population), as previously described (Shen et al. 2021). Individual colonies were identified by the distinct morphology and red color, separating these erythroid cells from other hematopoietic colonies. Colony cells were selected 14 days after plating.

### Erythroid differentiation

HSPCs were cultured at 37 °C and 5% CO in phase I erythroid differentiation medium (days 0-7), followed by phase II erythroid differentiation medium (days 7-12), and finally phase III erythroid differentiation medium (days 12-21). Briefly, erythroid base medium is composed of IMDM (Life Technologies) with human holo-transferrin (200 μg/mL), recombinant human insulin (10 μg/mL), heparin (3 IU/mL), human AB plasma (2%), human AB serum (3%), and penicillin/streptomycin (1%). Phase I medium consists of erythroid base medium supplemented with three cytokines, erythropoietin (EPO, 3 IU/mL), stem cell factor (SCF, 10 ng/mL), and interleukin-3 (IL-3, 1 ng/mL). Phase II medium is erythroid base medium with two cytokines, erythropoietin (EPO, 3 IU/mL), and stem cell factor (SCF, 10 ng/mL). And phase III medium consists of erythroid base medium plus erythropoietin (EPO, 3 IU/mL), and an additional 1 mg/mL of human holo-transferrin. During phase I and II cells were maintained at a concentration of 10^5^-10^6^ cells/mL, and 1-5×10^6^ cells/mL during phase III. Key characteristic features of differentiating HSPCs and erythroblast cells were confirmed using immunophenotyping of canonical cell-surface markers of early and late erythropoiesis (CD34, CD36, CD71 and CD235a). Primary erythroid cells (1-5 × 10^4^ cells) were incubated with antibodies for 15 min in the dark at room temperature. Cells were then washed twice with PBS/0.1% BSA. Flow cytometry was conducted on an Accuri C6 instrument and all data was analyzed using FlowJo software (v.10.3). Furthermore, forward and side scatter (FSC and SSC) gating were used to identify live cells and exclude doublets (all experiments showed comparable results, including edited cells). In addition, characteristic morphological changes were confirmed using hematoxylin and eosin staining of cell smears.

### CRISPR/Cas9 genome editing in HSPCs

CRISPR/Cas 9 genome editing in an isogenic setting of human HSPCs from healthy donors was used to create a genetic perturbation of the *SUPT5H* gene, relying on non-homologous end joining to mimic patient mutations (Shen et al. 2021). Guides (Key Resources Table) were generated as oligonucleotides and designed based on the genetic variations found in our patient cohort using Synthego CRISPR design tool. The Cas9 and gRNA complex (ribonucleoprotein, RNP) were delivered into HSPCs using the Amaxa Nucleofector System (program DZ-100). Briefly, gRNA (5 μL of 120 pmol) and Cas9 protein (105 pmol, Integrated DNA Technologies) were mixed to form RNP complexes and delivered in HSPCs (20 μL of 2–4×10^5^) using the P3 Primary Cell 4D Nucleofector X Kit S (Lonza, following the manufacturer’s instructions). Cells were then transferred to phase I culture medium for 24 h and either seeded in MethoCult medium (for colony-forming assays) or kept in culture (for bulk assays). Edited colonies and bulk cells were isolated (after 2 weeks or 4 days, respectively) to obtain both the gene expression (through mRNA analysis), and the specific genome-editing genotype in bulk cells or in each colony (using CRISPR amplicon sequencing). DNA and RNA were simultaneously isolated using the ALLPrep DNA/RNA Micro Kit (Qiagen), and primer sequences used for CRISPR sequencing of gRNA editing efficiency are shown in the Key Resources Table and Table S9.

### Immunoblot analysis

Immunoblotting was performed as previously described (Mayer and Churchman 2016). Briefly, cells were lysed in RIPA buffer (Santa Cruz Biotechnology) for RNA-seq samples, or nuclear lysis buffer for NET-seq samples on days 8 and/or 11 of differentiation to confirm that cells maintained their SPT5 KD during erythropoiesis. Samples were analyzed using antibodies against SPT5 (Millipore, MABE1803), Polymerase II RPB1 (BioLegend, 8WG16), and H3 (Cell Signaling, 9715), and stained with LI-COR secondary antibodies. Blots were imaged on the ChemiDoc Imaging System (Bio-Rad). Quantified blotting intensities were analyzed using Bio-Rad Image Lab Software, normalized to the loading control (H3) and AAVS1 controls, and plotted in R.

### NET-seq, RNA-seq, and Nanostring direct mRNA detection

NET-seq was performed as previously described (Martell et al. 2021; Mayer and Churchman 2016; Mayer et al. 2015). Due to the changing cell types, NET-seq cellular fractionation efficiency was measured by western blot and subsamples of cultured HSPCs showed that ≥95% of elongating Pol II was captured in the chromatin fraction. A *Drosophila* spike-in control was added to each NET-seq sample for normalization (Table S10). Gene expression profiles between donors were highly correlated and much of the observed variability between biological replicates can be attributed to different collection days (i.e., the differentiation trajectory), or treatment (e.g., DMSO control vs DRB), or gRNA target (i.e., AAVS1 control vs SPT5), demonstrating high reproducibility between biological replicates. For RNA-seq and Nanostring direct mRNA detection of edited cells in culture, total RNA was extracted from whole cells using Qiazol reagent (Qiagen), and libraries were prepared according to the SMART-Seq v4 PLUS Kit (Takara). For RNA-seq and Nanostring direct mRNA detection of CRISPR colonies, DNA and RNA were simultaneously isolated using the ALLPrep DNA/RNA Micro Kit (Qiagen). Much of the observed variability between biological replicates can be attributed to the CRISPR guide RNA target (SPT5 vs AAVS1 control) and different collection days, demonstrating high reproducibility between biological replicates. For Nanostring direct mRNA detection, RNA purifying kit (Zymo) and probes were designed by Nanostring (Key Resources Table and Table S11) and used to directly detect mRNA levels.

### Cell cycle and proliferation

EdU labeling for cell cycle dynamics was performed on 5–10×10^4^ cells using the Click-iT EdU kit (Thermo Fisher) following the manufacturer’s instructions for the *Flow Cytometry Cell Proliferation Assay*. Dual EdU and BrdU labeling for cell cycle kinetic determination was performed on 2–5×10^5^ cells as previously described following the *Basic Protocol* (Bradford and Clarke 2011). The length of the S phase (Ts) was calculated as previously described (Martynoga et al. 2005). Briefly, Ts = Ti/(P_L_/P_S_), where Ts = S phase length, Ti = EdU (but not BrdU) labeling time, Ps = Portion of cells in S phase (EdU+/BrdU+), and P_L_ = Portion of cells that have left S phase (EdU+/BrdU-). Cell proliferation was measured using a luminescent assay (CellTiter-Glo™, Promega), a Countess cell counter, and/or manually with a hemocytometer.

### Inhibitor treatments

HSPCs were treated on day 8 of erythroid differentiation with SPT5 small-molecule inhibitors (SPIs) as well as various transcription inhibitors (DRB, triptolide, and flavopiridol). For all treatment conditions, cell viability was measured using a luminescent assay (CellTiter-Glo™, Promega) to verify a healthy cell population. Briefly, HSPCs were treated with SPIs for 2, 8, and 24 hrs at concentrations ranging from 0-50 uM and transcription inhibitor treatments were for 15, 30, and 60 minutes at varying concentrations (0-600 nM for flavopiridol, 0-10 uM for triptolide, and 0-200 uM for DRB). Samples were collected and processed according to the above-described assays and most of the observed variability between biological replicates can be attributed to the inhibitor treatment (e.g., DMSO control vs DRB), demonstrating high reproducibility between biological replicates.

## QUANTIFICATION AND STATISTICAL ANALYSIS

### Analysis of sequencing data

Differentially expressed genes from RNA-seq data were identified by pairwise analysis with DESeq2 (Love, Huber, and Anders 2014) at an adjusted p value of < 0.05. NET-seq data was aligned and processed as previously described (Martell et al. 2021; Mayer and Churchman 2016; Mayer et al. 2015). For NET-seq expression analysis on edited or inhibitor treated cells, gene counts were spike-in normalized. For NET-seq expression analysis during erythropoiesis, changes in expression over the course of differentiation were identified by a likelihood ratio test using DESeq2 at an adjusted p value of < 0.05 and a base mean count >10. NET-seq data analysis during normal erythropoiesis was also analyzed using spike-in normalized counts and results were consistent with TPM normalized counts; however due to varying *Drosophila* spike-in percentages (Table S10) we chose to only show the TPM normalized data. Counts for expression analysis were obtained by running featureCounts (Liao, Smyth, and Shi 2014) on processed bam files. Expression analysis of *HBA* used multi-mapped reads with fractional counts. The *HBB* and *HBA* LCR enhancer regions were defined by the NCBI GRCh38.p14 Primary Assembly. Read counts for IGV traces were calculated using deepTools (Ramírez et al. 2016) and TPM normalized. Data significance was analyzed by Student’s t-test. In all figures, data are represented by mean ± SEM from 2 to 3 donors, as noted in the figure legends. PCA was performed on the top 1,000 most variable genes. For heatmaps, k-means clustering was performed on z-score normalized values using the pheatmap package in R. The elbow method was used to determine the number of clusters. GO analysis on all heatmap clusters was performed using clusterProfiler (p adjusted < 0.05) and REVIGO (Supek et al. 2011) was used to get the top representative GO terms for the RNA-seq data. For Nanostring analysis, mRNA measurements were analyzed using the nSolver Software and normalized to the control probe (Key Resources Table).

### Pausing index analysis of NET-seq data

We used previously established standards to calculate pausing indices (PIs) and to set appropriate thresholds for including genes where PIs can be reliably measured (Adelman and Lis 2012). Specifically, PIs were calculated as the ratio of normalized counts in the promoter window (−30 bp to +300 bp around the TSS) over the gene body density (+700 to TTS) for genes greater than 1.5 kb in length. Counts were normalized by the length of the promoter window or gene body length. Counts were obtained for protein coding using featureCounts (Liao, Smyth, and Shi 2014) and TPM normalized (for the timepoints during erythropoiesis) or spike-in normalized (for edited or inhibitor treated cells). For the timepoints during erythropoiesis, we analyzed genes that had a normalized promoter region signal above 3 and a PI > 4 for at least one time point during erythroid differentiation to ensure that each gene is paused at some point during differentiation. To include genes which are paused but not expressed during erythroid differentiation, a pseudocount was added to the gene body density count. For edited or inhibitor treated cells, genes were selected that had a normalized promoter region signal above 3 and a PI > 2 for at least one condition, such that each gene experiences measurable pausing after treatment or control.

### Metagene analysis of NET-seq data

For edited and inhibitor treated cells, genes with normalized counts >= 5 were selected for analysis. Biological replicates were averaged together, and genes were normalized by their average signal within +/- 2kb of the TSS. The mean NET-seq signal at each site surrounding the TSS was then calculated and plotted.

## Supplementary Figure Legend

**Figure S1. Pol II promoter-proximal pausing during human erythroid differentiation**. (A) Representative western blot showing the quantitative purification of elongating RNA polymerase II by cell fractionation. Subcellular fractions were obtained from ∼1×10^6 human CD34+ cells; antibodies directed against Pol II CTD Ser2-phosphorylated and GAPDH. (B) Quantification of western blots from cell fractionations of daily subsamples during human erythropoiesis; image quantification was obtained using ImageJ. (C) Hematoxylin and eosin staining of cell smears confirming characteristic morphological changes during erythropoiesis, images taken with 630X oil immersion. (D) FACS of canonical cell-surface markers (CD71 and CD235a) following erythropoiesis of healthy donors in culture. (E) Pearson correlation heatmap of normalized NET-seq expression (log transformed) across days of differentiation for two donors. (F) Heatmap of differentially expressed genes from likelihood ratio test (n= 4,533, adjusted p-value <0.05) using k-means clustering (resulting in 3 clusters) across erythroid differentiation. Values are z-score of per day average normalized expression; n = 2 donors. (G) Enriched Gene Ontology biological processes for all three clusters of differentially expressed genes; the top five representative processes are shown and their -log10(p adjusted value). (H) Bar plot of normalized NET-seq globin expression during erythropoiesis; n = 2 donors, data are represented as mean ± SEM.

**Figure S2. Pol II promoter-proximal pause release is hindered in human hematopoietic cells upon perturbation of *SUPT5H***. (A) Editing efficiency of gRNAs evaluated by CRISPR sequencing in erythroid colonies for one control guide (*AAVS1*) and two guides targeting *SUPT5H* (results were consistent between the two *SUPT5H*-targeted samples); data are represented as mean ± SEM, n > 10 independent experiments for each gRNA. (B) (Left) Western blot of Pol II (RPB1) levels; erythroid cells on day 11 in culture treated with gRNAs targeting the *SUPT5H* gene or an *AAVS1* control. H3 serves as a loading control. (Right) Western blot quantification of Pol II knockdown. Signal was normalized to H3 and *AAVS1*; data are represented as the mean, n= 3 donors, t-test. (C) Bar plots for erythroid colonies showing the ratio of *HBB* relative to *HBA* calculated from direct mRNA levels for all gRNAs; n > 10 independent experiments for each gRNA. (D) Average globin mRNA levels for erythroid colonies, all globins shown (except for *HBA*) are regulated by the *HBB* LCR; n > 10 independent experiments for each gRNA. (E) Principal component analysis of NET-seq expression on day 8 of erythropoiesis, colored by gRNA target; n= 3 donors. (F) Ratio of *HBB* relative to *HBA* for NET-seq data; data are represented as mean ± SEM, n= 3 donors, t-test. (G) Pausing indices of *SUPT5H*-edited cells relative to *AAVS1* control for *HBB* and *HBA*; data are represented as mean ± SEM, t-test. (H) Enhancer reads in hypersensitive sites of the *HBB* LCR and (I) *HBA* LCR, comparing *SUPT5H*-edited cells to *AAVS1* control; data are represented as mean ± SEM, n = 3 donors, t-test. P-values: *: p<0.05; **: p<0.01; ***: p<0.001

**Figure S3. Perturbation of *SUPT5H* delays erythroid gene expression programs**. (A) Principal component analysis plot of RNA-seq expression on erythroid colonies and colored by gRNA target, *AAVS1* control or *SUPT5H*; n = 3 independent experiments. (B) Principal component analysis plot of RNA-seq expression on edited erythroid culture cells (*AAVS1* control or *SUPT5H*) colored by time point; n = 3 donors. (C) Gene Ontology biological processes for differentially expressed genes in bulk edited cells (comparing *AAVS1* and *SUPT5H*) on day 4 of erythroid differentiation; the top ten enriched representative processes are shown and their - log_10_(p adjusted value). (D) Average *SUPT5H* expression relative to *AAVS1* control at different timepoints during erythroid differentiation; t-test. (E) Average *HBB* and *HBA* mRNA levels on day 4 of erythroid differentiation in *AAVS1* and *SUPT5H*-edited cells; t-test. P-values: *: p<0.05; **: p<0.01; ***: p<0.001

**Figure S4. Reduced levels of *SUPT5H* result in a stage-specific delay in erythroid differentiation**. (A) Representative FACS plot of erythroid terminal differentiation markers CD36 and GPA; plot shows day 21 of the *SUPT5H*-edited cells and the *AAVS1* control (B) Quantification of decrease in CD36 as cells progress in erythroid terminal differentiation, showing no significant difference between *SUPT5H*-edited cells and the *AAVS1* control; measured on days 16 and 21; data are represented as mean ± SEM; n = 3 donors, t-test. (C) Representative FACS plot of erythroid terminal differentiation markers CD71 and GPA; plot shows day 21 of the *SUPT5H*-edited cells and the *AAVS1* control. (D) Quantification of decrease in CD71 as cells progress in erythroid terminal differentiation, showing no significant difference between *SUPT5H*-edited cells and the *AAVS1* control; measured on days 16 and 21; data are represented as mean ± SEM; n = 3 donors, t-test. (E) Hematoxylin and eosin staining of cell smears confirming characteristic morphological changes and showing no significant difference between *SUPT5H*-edited cells and the *AAVS1* control; imaged on days 10, 16 and 21 in culture, images taken with 630X oil immersion.

**Figure S5. Perturbing *SUPT5H* leads to stage-specific changes in the cell cycle**. (A) Line plot showing increased cell proliferation in *SUPT5H*-edited cells measured by ATP bioluminescence during *ex vivo* erythroid differentiation from days 1 to 11 in culture. *SUPT5H*-edited cell signal and *AAVS1* control signal are relative to day 1 and nucleofection with gRNA was on day 1 in culture; n = 3 donors. (B) Average cell number in *SUPT5H*-edited cells relative to *AAVS1* control for two *SUPT5H* target gRNAs. Plot shows day 8 of erythroid differentiation in culture; n = 3 donors. (C) Average cell number in *SUPT5H*-edited cells relative to *AAVS1* control for five different donors. Plot shows day 8 of erythroid differentiation in culture; n = 2 independent experiments. (D) Representative FACS plot of EdU labeling to measure cell cycle populations; plot shows days 2 to 6 of erythroid differentiation in culture, lower left quadrant shows the G0/G1 population, lower right quadrant shows the G2/M population, top quadrant shows the S population; quadrant percentages are mean ± SEM, n= 3 donors.

**Figure S6. Transcription inhibitors disrupt globin balance and recapitulate the β-thalassemia phenotype**. (A) Cell viability was measured by ATP luminescence for transcription inhibitor treated cells; viability is relative to the DMSO control, n = 3 donors. (B) Cell viability measured by ATP luminescence for SPT5 small-molecule inhibitor (SPI) treated cells; viability is relative to the DMSO control, cells were treated for 2 hours on day 8 of erythroid differentiation, n = 3 donors. (C) Principal component analysis plot of NET-seq expression comparing the DMSO control and DRB, n = 2 donors. (D) Principal component analysis plot of NET-seq expression comparing the DMSO control and SPI-18, n = 2 donors. (E) Average sense-strand normalized NET-seq signal around the transcription start site for genes comparing DMSO and SPI-18; n = 2 donors. (F) Ratio of *HBB* relative to *HBA* for NET-seq data comparing the DMSO control and transcription inhibitor treated cells; data are represented as mean ± SEM, n = 2 donors, t-test. (G) Ratio of *HBB* relative to *HBA* for NET-seq data comparing the DMSO control and SPI treated cells; data are represented as mean ± SEM, n = 2 donors, t-test. (H) Ratio of *HBB* relative to *HBA* calculated from direct mRNA levels using Nanostring comparing the DMSO control and transcription inhibitor treated cells; cells treated on day 8 of erythroid differentiation for 15, 30, or 60 minutes with a range of inhibitor concentrations; data are represented as the mean, t-test, n = 3 donors. P-values: *: p<0.05; **: p<0.01; ***: p<0.001

## Supplementary Table Legend

**Table S1. Hematological parameters and pathogenic variants of individuals with *SUPT5H* mutations**. Thalassemia screening of patients in a rare blood disorder cohort and the pathogenic variants in *SUPT5H* harbored by these patients. The Hb, MCV and MCH in *SUPT5H* carriers is markedly reduced and HbA 2 is elevated, consistent with a β-thalassemia trait phenotype.

**Table S2. Pausing index during erythropoiesis**. Pausing indices of protein-coding genes that have a normalized promoter region signal above 3 and a PI > 4 for at least one time point during erythroid differentiation to ensure that each gene is paused at some point during differentiation.

**Table S3. CRISPR colony gRNA editing and Nanostring data**. Genome-edited genotype obtained by CRISPR amplicon sequencing and Nanostring direct mRNA measurements normalized to the control probe for edited erythroid colonies.

**Table S4: Pausing index for *AAVS1* and *SUPT5H*-edited cells**. Pausing indices of protein-coding genes that have a normalized promoter region signal above 3 and a PI > 2 for at least one condition. Cells were collected on day 8 of erythroid differentiation

**Table S5. Pausing index for cells treated with SPT5 small-molecule inhibitors**. Pausing indices of protein-coding genes that have a normalized promoter region signal above 3 and a PI > 2 for at least one condition. Cells were treated with inhibitors for 2 hours on day 8 of erythroid differentiation.

**Table S6. Pausing index for cells treated with transcription inhibitors**. Pausing indices of protein-coding genes that have a normalized promoter region signal above 3 and a PI > 2 for at least one condition. Cells were treated with inhibitors for 30 minutes on day 8 of erythroid differentiation.

**Table S7. SPT5 small-molecule inhibitor Nanostring data**. Nanostring direct mRNA measurements normalized to the control probe from cells treated with SPT5 small-molecule inhibitors on day 8 of erythroid differentiation.

**Table S8. Transcription inhibitor Nanostring data**. Nanostring direct mRNA measurements normalized to the control probe from cells treated with transcription inhibitors on day 8 of erythroid differentiation.

**Table S9. CRISPR sequencing oligos**. Primer sequences used for CRISPR amplicon sequencing of genome-edited cells.

**Table S10. Sequencing metadata for NET-seq samples**. Metadata on sample treatment, day in erythroid differentiation culture, de-identified donor number, and *Drosophila* spike-in percentage for NET-seq sequencing libraries.

**Table S11. Nanostring direct mRNA detection probes**. Primer sequences used for Nanostring probes to directly detect mRNA levels.

