## Supplementary figures and images for "RNA Polymerase II pausing temporally coordinates cell cycle progression and erythroid differentiation"

### Supplemental Figure

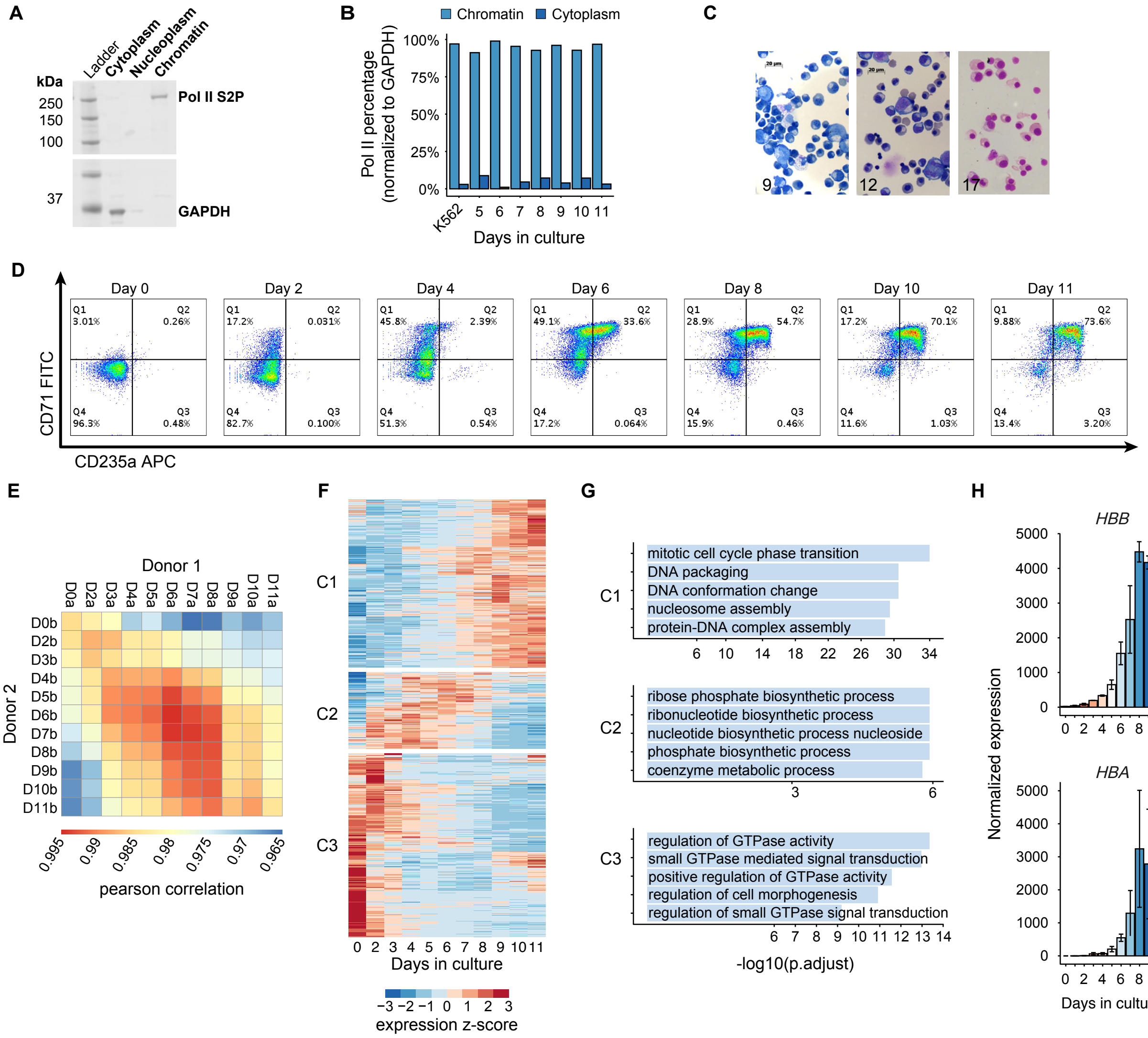

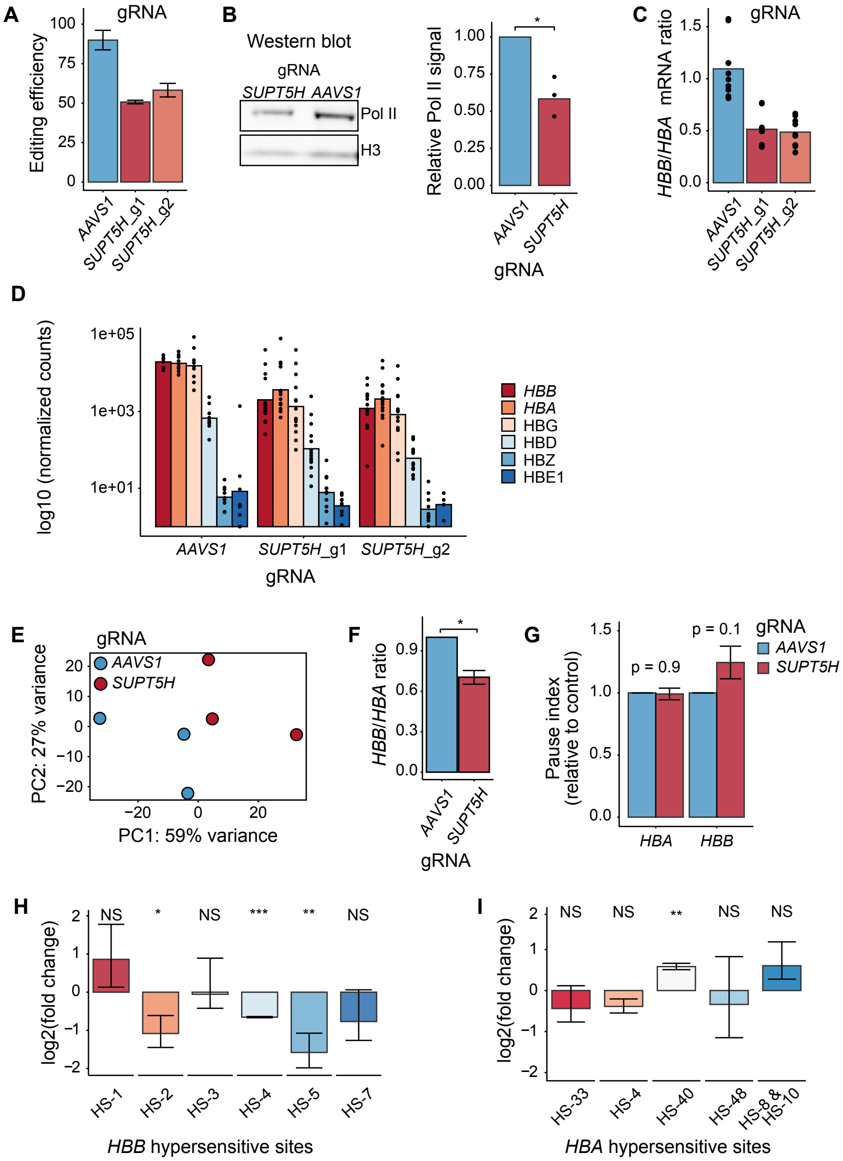

#### Figure S3

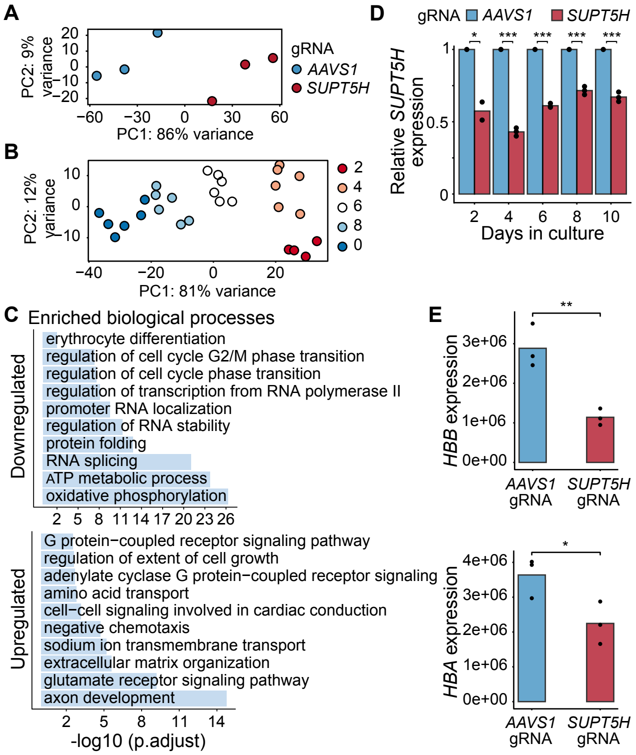


#### Figure S4

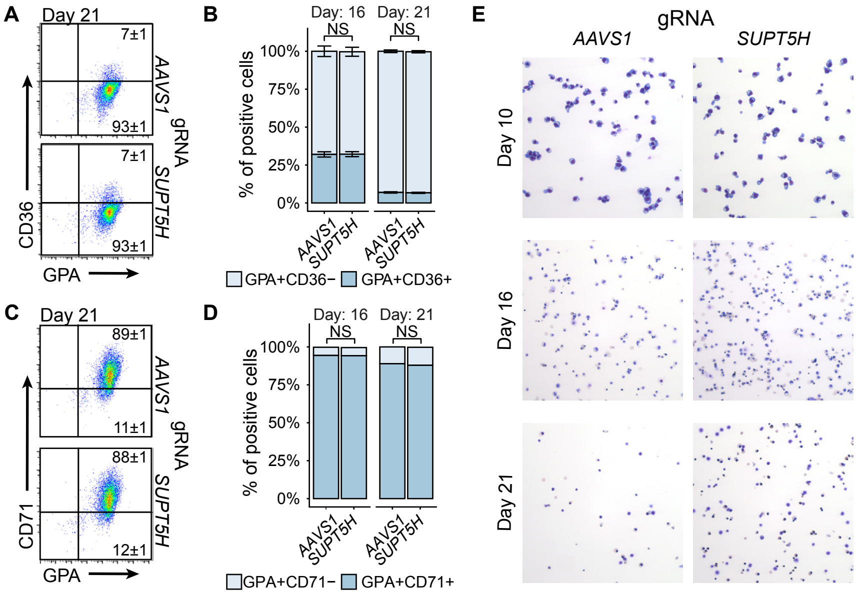

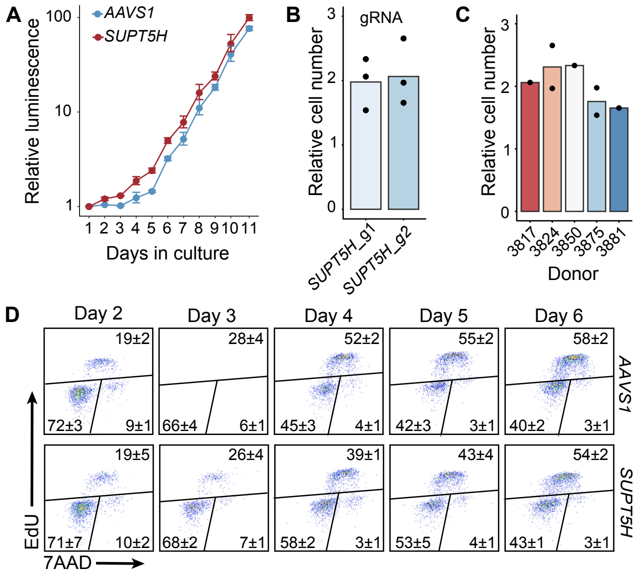

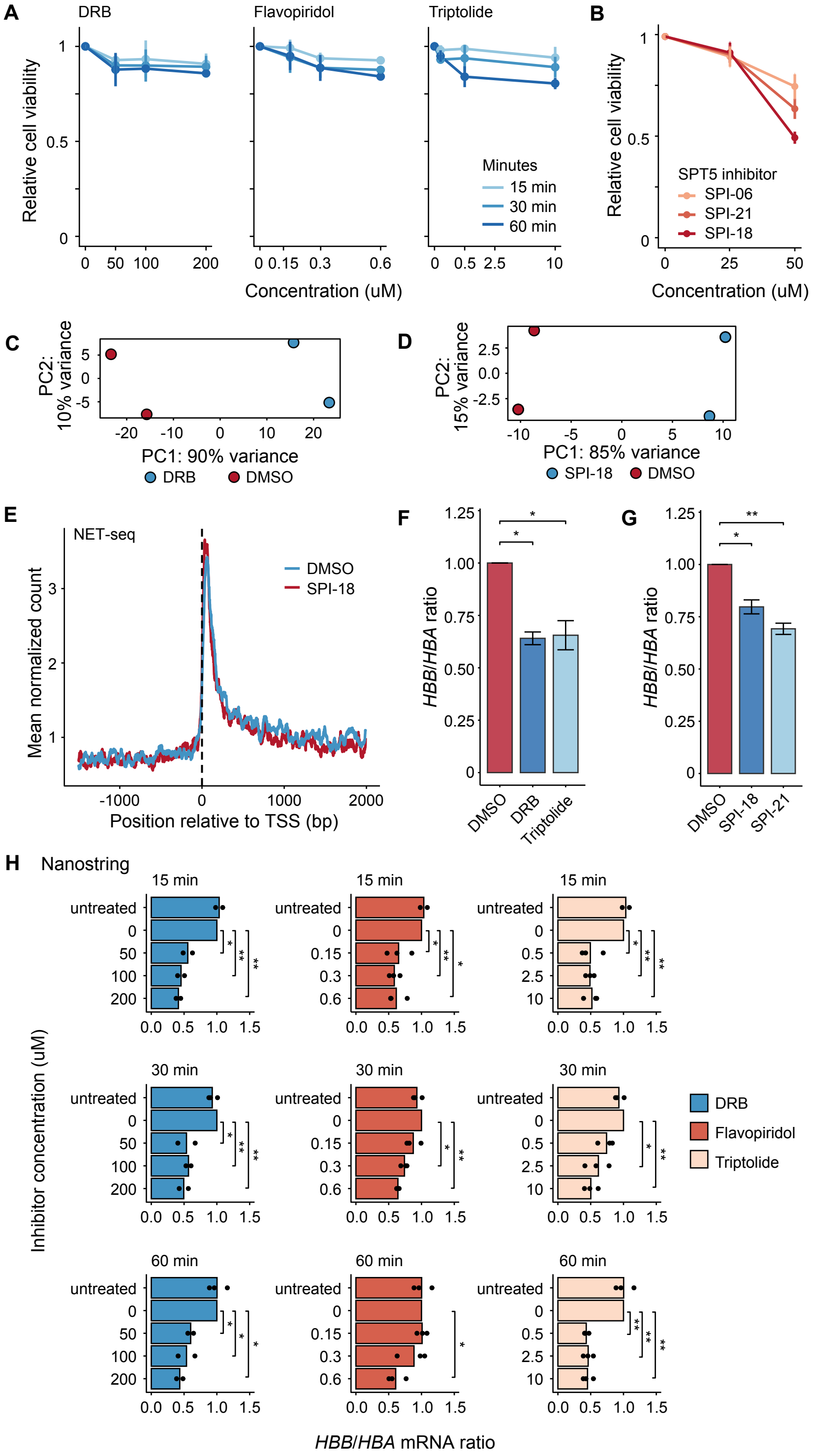
